# The effect of neuromuscular blockade on EEG-based measures of awareness

**DOI:** 10.1101/2025.07.11.25331259

**Authors:** Sebastian Halder, Bjørn E. Juel, Kenneth J. Pope, Andrew Hardy, John O. Willoughby, Johan F. Storm

## Abstract

**Background:** Both basic and clinical consciousness research aims to find objective measures that reliably distinguish conscious from unconscious brain states. Electroencephalogram (EEG) measures are widely used, although they may be contaminated by electrical signals from muscles.

**Methods:** To assess this source of error, we investigated the impact of neuromuscular blockade (NMB) on proposed measures of consciousness (spectral slope, Lempel-Ziv complexity (LZc), connectivity, alpha peak frequency, power in canonical EEG frequency bands) computed from spontaneous high-density EEG recorded from six healthy volunteers in three different conditions: (1) awake-unparalysed, (2) awake-paralysed caused by neuromuscular blocking agent (NMBA), and (3) sedated-paralysed (sedated with propofol, paralysed by NMBA, (un)consciousness non-confirmable).

**Results:** The markers we investigated distinguished awake-unparalysed states from sedated-paralysed with close to perfect accuracy. Our analysis revealed a serious failure of all measures, except alpha power, to recognise awake-paralysed, without sedation, as an aware state. Errors ranged from 19% of awake-paralysed time segments predicted as unaware (using spectral slope) to 100% (using LZc). Using alpha power, only 1% of all awake-paralysed segments were misclassified. Critically, the awake-paralysed is the state that is important to detect in sedated-paralysed patients, to prevent the experience of accidental awareness during general anaesthesia (AAGA).

**Conclusions:** This study clearly demonstrates that many EEG-based measures fail to recognise awareness in awake-paralysed subjects, by using a unique high-density EEG data set. Alpha power was determined to be the most robust measure to detect AAGA, but this may not generalise to all types of general anaesthetic agents.

## 1 Introduction

Objective measures of consciousness should reliably distinguish conscious from unconscious brain states. Developing such measures involves comparing wakefulness with states of apparent unconsciousness, such as deep sleep or general anaesthesia. However, this approach implicitly equates unresponsiveness with unconsciousness.^1,2^ In particular, the paralysed state induced by neuromuscular blocking agents (NMBAs), in which a patient appears unresponsive, may be mistaken for unconsciousness.^3^ Thus, an objective measure of consciousness must go beyond distinguishing responsive-awake from unresponsive-unconscious states. It must specifically be able to differentiate between a state that is conscious but unresponsive (such as accidental awareness during general anaesthesia (AAGA)) and a state that is genuinely unconscious and unresponsive (such as successful general anaesthesia).

Neuromuscular blockade (NMB) is used in 62% of general anaesthetic procedures, for easing intubation and optimising patient positioning.^4,5,6^ NMBAs often have a longer duration of effect than anaesthetics,^7^ leading to the risk that patients experience AAGA. AAGA is estimated to affect 1 in ∼900 patients^8^ with young adults and females at increased risk,^9^ and can be highly traumatic.^10^

Commonly used indices such as bispectral index (BIS)^11,12^ and GM Entropy monitors incorporate electromyogram (EMG) activity, which may be informative in unparalysed patients. However, during general anaesthesia with NMB, EMG is intentionally suppressed, limiting the utility for detecting AAGA.^13,14,15^ From the perspective of detecting awareness specifically in paralysed patients, the absence of EMG represents an issue that must be allowed for — dependence on EMG signals represents a fundamental limitation for any method of AAGA detection in paralysed patients.

Over the past decades, hundreds of electroencephalogram (EEG)-based measures have been proposed as ”measures of consciousness”. Gathering enough data from surgical patients to evaluate measures of awareness is challenging because AAGA rarely occurs and explicit recollection of the event may be impaired.^8^ Thus, many measures have largely been tested in controlled laboratory environments, and it remains unclear whether they are sensitive to EEG changes induced by NMBA, i.e. to EMG contamination.^16^ Thus, the reliability of these EEG-based measures in detecting AAGA, and thus their ability to detect awareness as opposed to unresponsiveness, remains questionable.

Characterizing how different EEG-based measures respond during NMB provides important information about their clinical utility in settings where NMBAs are used. We therefore quantified the differential responses of state-of-the-art measures based on Lempel-Ziv complexity (LZc),^17^ periodic and aperiodic components of the power spectrum,^18,19^ and functional connectivity^20^ during paralysis with NM-

BAs. Ideally, these measures would remain stable between the awake-unparalysed and awake-paralysed states, both conscious conditions, while showing a clear difference between these awake states and the paralysed-sedated state. This hypothesis reflects the fundamental expectation that valid consciousness measures track awareness itself, independent of motor output or muscle activity, which is critical for detecting awareness during neuromuscular blockade. If these measures fail to make this distinction, they cannot reliably detect AAGA or consciousness more broadly. To test this hypothesis, we analysed a unique EEG dataset recorded from healthy volunteers in three distinct states: (1) awake-unparalysed, (2) awake-paralysed, and (3) sedated-paralysed with propofol and NMB.

## 2 Methods

The study was approved by the Southern Adelaide Clinical Human Research Ethics Committee. Participants gave written, informed consent for the procedures.

### 2.1 Terminology

Terms such as “consciousness” and “awareness” have been used in various ways, sometimes causing confusion. As explained in the Supplement, we here use the terms “conscious”/“consciousness” for the state/ability to experience anything, including dreaming, even when the subject is unable to respond. “Awareness” denotes here the ability to experience current sensory input from the body and world, but not dreaming, while “responsiveness” is the ability to respond verbally or non-verbally to stimuli.^2^ To denote the three states that we compare in this study, we use the following short terms: (1) “awake-unparalysed” (responsive, aware, conscious), (2) “awake-paralysed” (caused by NMBA only, responsive with isolated forearm technique (IFT), aware, conscious), and (3) “sedated-paralysed” (caused by NMBA plus propofol, overtly unresponsive, subjectively unaware and unconscious, with no reported memories from the sedated period but the level of (un)consciousness could not be objectively verified due to NMB and release of IFT).

### 2.2 Data

EEG was recorded with a 115 channels Neuroscan EEG amplifier (Compumedics, Victoria, Australia), at 5000 Hz with a 1250 Hz low-pass filter, a 128-channel cap with Ag-AgCl electrodes (Easy Cap GmbH, Germany), reference placed on the left ear, impedances were below 5*k*Ω, electrooculogram, electro-cardiogram, respiration and EMG sensors. The sedation recordings, though collected alongside earlier work,^21,22,23^ remained unanalysed until this study. The current authors initiated re-examination of the full dataset to address previously unexplored questions regarding effects of NMB on EEG signatures of consciousness.

### 2.3 Experimental procedure

EEG was recorded from n=6 volunteers (one female) in three states (always with with eyes closed, see above). Participants received a dose of glycopyrrolate (see **Table** 1 for all dosages) and the laryngeal mask airway (LMA) was inserted (awake-unparalysed, see **Figure 1**). The IFT^3^ was applied and NMBA was administered. Paralysis was confirmed via EMG (awake-paralysed). The IFT was deflated at the limit of ischaemic tolerance. From then on the participants were unable respond (and responsiveness could no longer be confirmed if present). Subsequently participants were sedated using Propofol. Propofol was administered using a Graseby 3500 Anaesthesia Pump incorporating Diprifusor TCI Version 2, which employs the Marsh pharmacokinetic model. All subjects received a machine-calculated bolus followed by an infusion titrated to maintain the target plasma concentration (**see Table 1**). The bolus doses were consistent with the Diprifusor used for deep sedation, not anaesthesia, and each bolus was followed by an infusion (sedated-paralysed). All six participants reported no conscious experience during this period when asked right after the experiment (which included train-of-four (TOF) testing supramaximal electrical stimulation of a peripheral nerve that is perceived as unpleasant, performed before administering the NMBA reversal agent). Nonetheless, neither unconsciousness nor unresponsiveness during sedated-paralysed could be objectively verified at the time of EEG recording. The aim of the sedation was to maintain tolerance of the LMA in a non-theatre environment until the neuromuscular blockade could be safely reversed. Four participants received a dose of fentanyl at the time of the propofol bolus (Participants 3 and 5 were opioid intolerant). Reversal of neuromuscular blockade was initiated only after TOF monitoring indicated the onset of spontaneous recovery, ensuring that the paralysis state was maintained during EEG acquisition in the sedation phase. Due to a protocol deviation Neostigmine was administered before recording the sedated state for Participant 5. Participants were asked to perform different mental tasks during the awake-unparalysed and awake-paralysed state which were not analysed in the present work (see Witham et al., 2008^23^).

**Figure 1.**
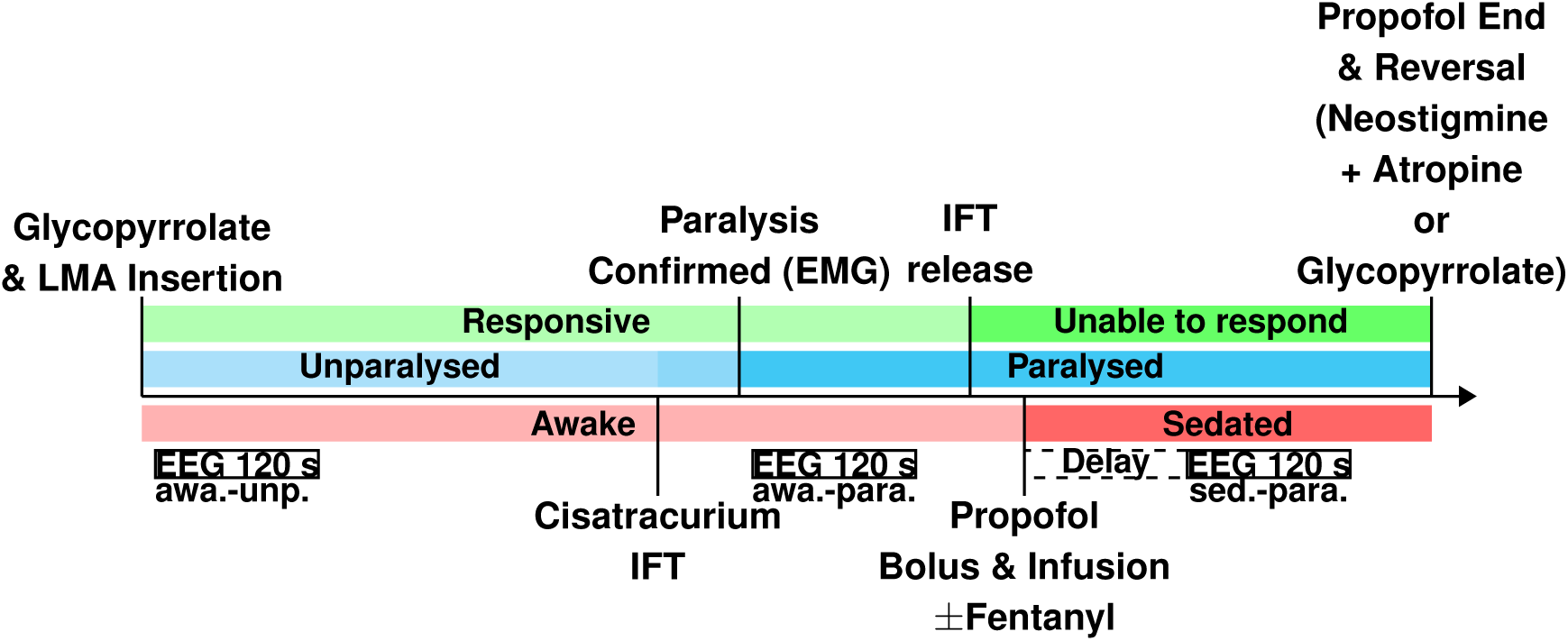
Timeline illustrating the sequence of key events and physiological states during the anaesthetic protocol with labels describing specific events (drug administrations, procedural steps, confirmations). Shaded rectangles indicate the occurrence of different states: ’Awake’ changed to ’Sedated’ after administration of propofol (red), ’Unparalysed’ changed to ’Paralysed’ after administration of the NMBA Cisatracurium (blue), ’Responsive’ changed to ’Unable to respond’ after IFT release (green). White boxes below each state rectangle indicate 120-second EEG segment used for the current analysis. Below each box we indicate which designation we use for this state in the text (awa.-unp.: awake-unparalysed, awa.-para: awake-paralysed, sed-para.: sedated-paralysed). Due to a protocol deviation Neostigmine was administered before recording the sedation EEG for Participant 5. Note that the sizes of the rectangles are not proportional to the duration in seconds.

**Table 1.** Summary of sedation and neuromuscular blockade protocol. The table includes subject demographics, propofol dosing parameters, and neuromuscular blockade management details. Target plasma concentration of propofol varied between experiments, with initial targets ranging from 2 mcg ml^−1^ to 4 mcg ml^−1^. Neostigmine (2.5 mg) was administered with either atropine (1.2 mg) or glycopyrrolate (0.4 mg) for reversal of neuromuscular blockade. Total propofol doses are reported in milligrams (mg), with ”NS” indicating this was ”not specified” in the records. Notes: ^1^Initial target plasma concentration of propofol set at 2 mcg ml^−1^; participant was persistently hypertensive during sedation phase. ^2^Participant 1 experienced variable target concentrations during infusion, as detailed in the table due to signs of clinically inadequate sedation. ^3^Range reflects progressively increased target concentrations during infusion for Participant 1.

| Part. | Age (range) | Sex | Weight (kg) | Target (mcg ml<sup>-1</sup>) | Bolus (mg) | Bolus (sec) | Infusion (mins) | Total (mg) | Fentanyl | Reversal |
| --- | --- | --- | --- | --- | --- | --- | --- | --- | --- | --- |
| 0 | 61-65 | M | 80 | 2<sup>1</sup> | 37 | 10 | 45 | NS | 100 µg | Neo. + Atrop. |
| 1 | 41-45 | M | 105 | Variable<sup>2</sup> | 48–240<sup>3</sup> | 14–21<sup>3</sup> | 40 | 1100 | 100 µg | Neo. + Atrop. |
| 2 | 71-75 | M | 95 | 4 | 87 | 26 | 30 | 550 | 100 µg | Neo. + Atrop. |
| 3 | 56-60 | M | 65 | 4 | 59 | 17 | 33 | NS | N/A | Neo. + Atrop. |
| 4 | 36-40 | M | 78 | 4 | 71 | 21 | 40 | 800 | 50 µg | Neo. + Atrop. |
| 5 | 46-50 | F | 68 | 4 | 62 | 18 | 22 | 200 | N/A | Neo. + Glyco. |

### 2.4 Pre-processing

All data analysis was performed using Python MNE.^24^ The original recording was downsampled to 250 Hz, re-referenced to REST,^25^ a reference-standardization approach that reduces volume conduction effects and distortions of topographic distributions, and high-pass filtered at 1 Hz. We extracted a 120 sec epoch from each state (see **Figure 1**) which were split into 5 sec epochs with 50% overlap (47 epochs per participant and condition).

### 2.5 Feature extraction

We extracted five sets of features. (1) Logarithmic *bandpower* from the canonical frequency bands using fitting oscillations & one-over-F (FOOOF)^19^ to parameterise the power spectra. (2) *Spectral slope/exponent* of the aperiodic component of the canonical bands, the full band (1-45 Hz), the low half (1-20 Hz) and the high half (20-40 Hz).^18^ (3) *Peak frequency* of each of the bands in Hz as determined with FOOOF. (4) *LZc* was computed for each epoch and channel on the time domain signal using single channel Lempel-Ziv Complexity.^17^ (5) *directed transfer function (DTF) (Outflow)*,^20^ was computed by estimating brain connectivity with DTF^26^ and computing the median of all outgoing connections for each electrode. In an exploratory analysis we expanded our original selection of features to a total of 55 measures derived from the metrics Casey et al.^27^ investigated. See supplementary material for details.

### 2.6 Classification

We evaluated how accurately we could predict the state of the participants using features from 5 sec EEG epochs using a linear support vector classifier (SVC). Training and testing was performed following a leave-one-participant-out scheme (five of the participants were used for training and feature selection, the remaining participant was for testing). We labelled each epoch as either ’aware’ (awake-unparalysed and awake-paralysed) or ’unaware’ (sedated-paralysed; please note discussion above in that we could not objectively verify if the participants were unconscious). Training and testing used two sets of classes. (1) *naïve:* the awake-paralysed data was excluded from training and testing. The ”naïve” approach resembles the standard way EEG measures of consciousness are tested—if they can reliably distinguish awake-unparalysed from sedated-paralysed, they are taken to work well—and we include it here to reproduce prior work. (2) *Two classes*: awake-paralysed data was excluded only from training but included in testing with the same label as the awake-unparalysed data (i.e. ’aware’). In the confusion matrices we indicate the true label of the awake-paralysed class resulting in a 2 × 3 matrix. ”Two classes” was the crucial test in this paper—measures should reliably classify participants under the effect of NMBAs as aware to be considered measures of awareness. Performance was evaluated using confusion matrices and *F*_1_-scores. See supplementary data for further details.

### 2.7 Statistical analysis

We assessed statistical significance using paired t-tests with Bonferoni correction (**Table S2**).

## 3 Results

For a measure to be clinically useful in detecting awareness during paralysis, it should show no significant difference between the two ’aware’ states (awake-unparalysed and awake-paralysed), but a clear difference between the ’aware’ states and the sedated-paralysed (’unaware’) state. This pattern would indicate that the measure may track awareness itself, rather than muscle activity. In terms of classification of individual epochs awake-unparalysed and awake-paralysed epochs should be labelled as ’aware’ and epochs recorded during sedated-paralysed should be classified as ’unaware’ (with the caveat that unconsciousness could not be objectively verified in this state in our study).

### 3.1 Effects of paralysis and sedation on power spectrum

Visual inspection of the group-averaged power spectra revealed that the awake-unparalysed and awake-paralysed conditions were highly similar below 15 Hz, with both states showing prominent oscillatory activity in the alpha range distributed across the scalp. In contrast, the sedated-paralysed state was characterised by a marked increase in overall power, particularly in the alpha band, which also appeared broader and more pronounced. Both the sedated and paralysed state showed a notable reduction in high-frequency (>15 Hz) power compared to the awake-unparalysed conditions. See **Figure 2**.

**Figure 2.**
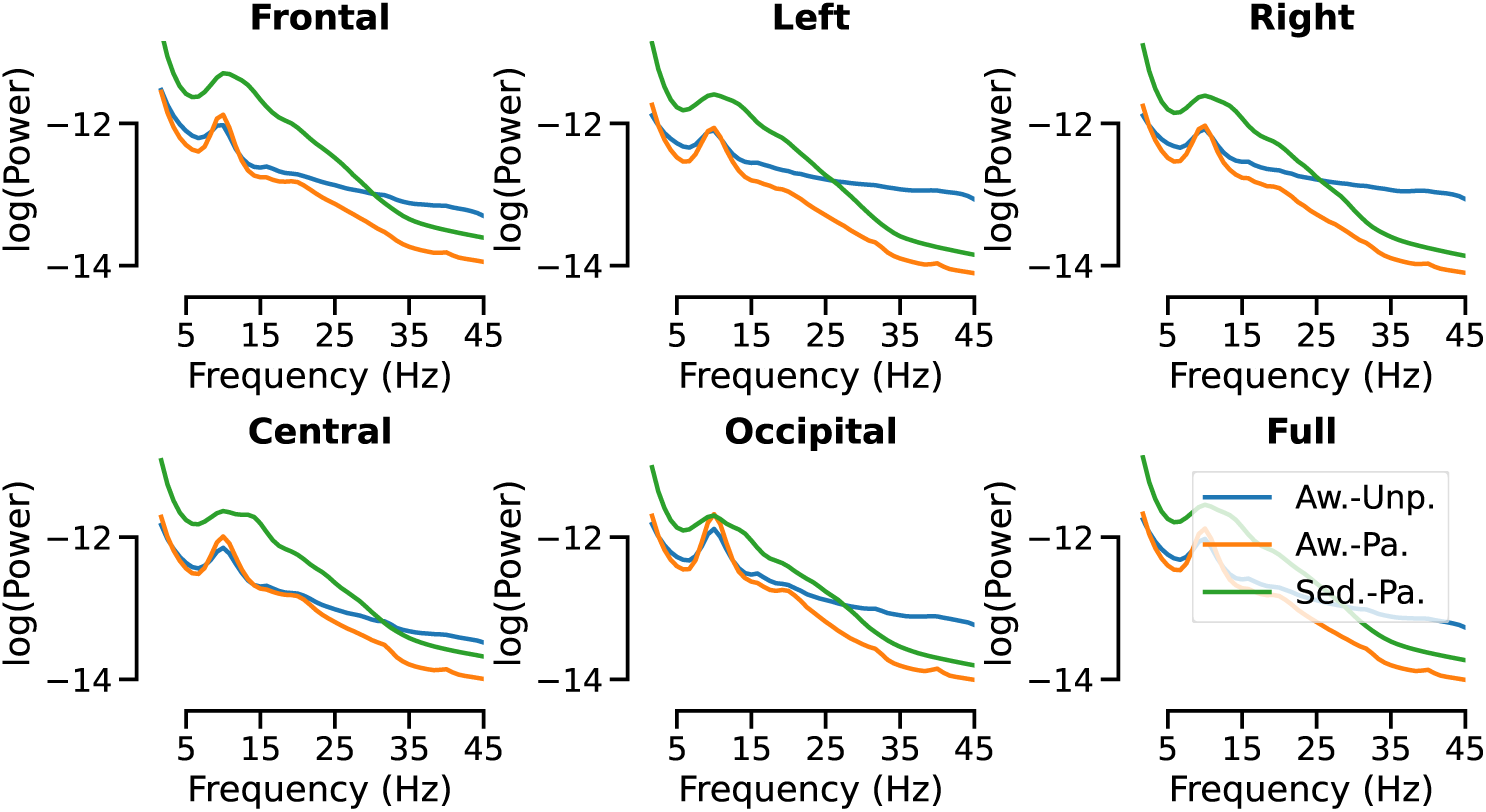
Power spectral densities visualised as line plots and averaged across all epochs and channels. The blue lines represent awake-unparalysed, the orange awake-paralysed and green sedated-paralysed. Regions of interest were visualised individually using separate lines per condition. The final plot shows the power spectrum computed using the full channel set. The changes between conditions varied depending on the frequency range and scalp location. For example, compared to normal wakefulness, gamma power decreased the most in peripheral locations, e.g. by 7.4% in the left region of interest (ROI) (from -12.9 in the awake-unparalysed to -13.9 in the awake-paralysed state). For comparison, the decrease in the same band in the central ROI was only 3.3% from awake-unparalysed to awake-paralysed. Meanwhile, the biggest increases in power were seen in the alpha range, from awake-unparalysed to sedated-paralysed, e.g. increasing by 7.9% in the frontal ROI (from -12.3 in awake-unparalysed to -11.4 in sedated-paralysed). The occipital ROI showed the smallest changes across the full range between all conditions, although the gamma power appeared to decrease from the awake-unparalysed to awake-paralysed state, and sedation led to a general steepening of the spectrum and less extreme alpha peak.

### 3.2 Effect of paralysis on measures of awareness

Awake-paralysed differed from awake-unparalysed for diversity, slope (high), theta and gamma, indicating dependence on EMG and they may not accurately detect AAGA. See **Figure 3**, and **Tables S1** and **S2** for details).

**Figure 3.**
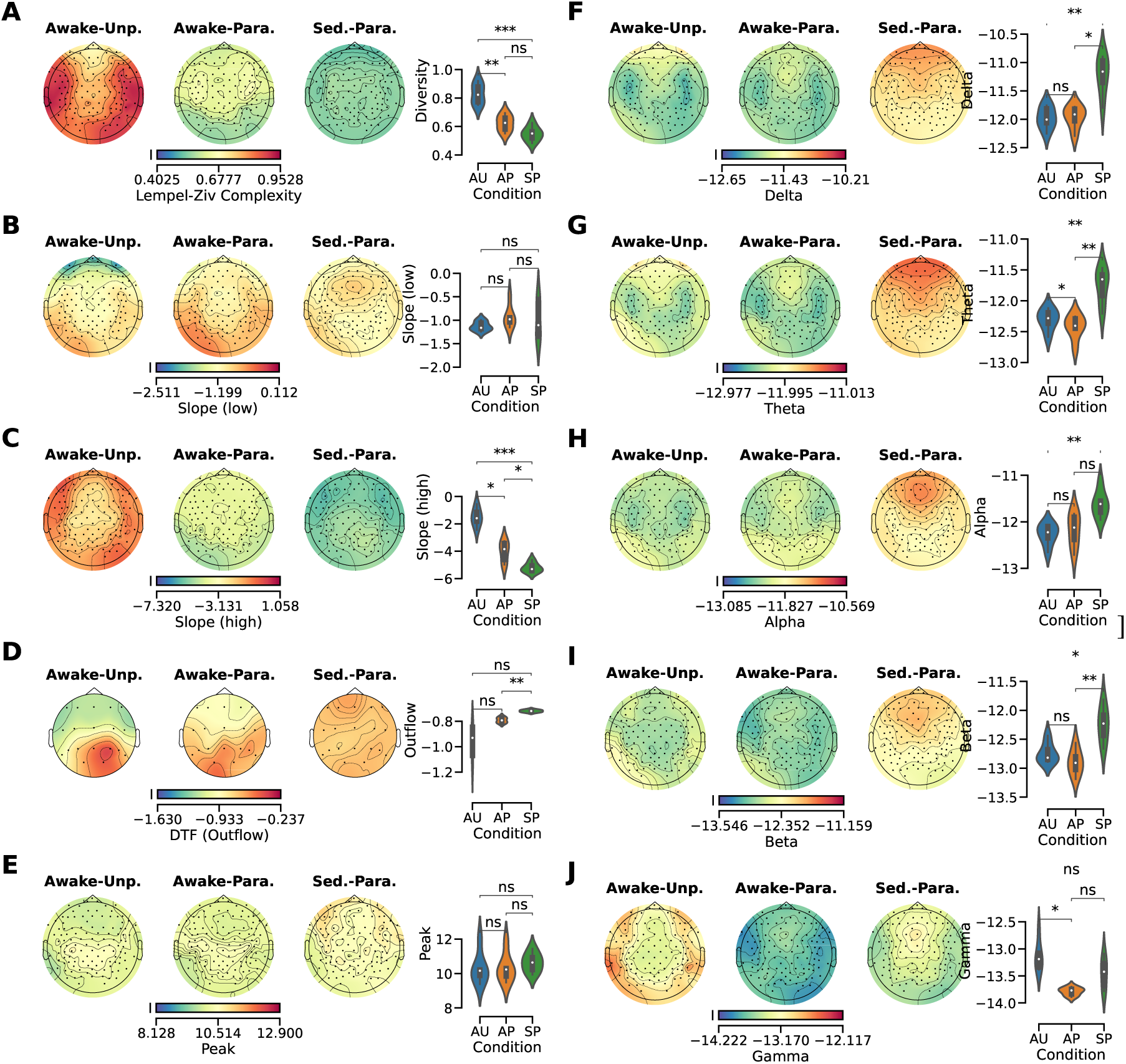
Difference of Lempel-Ziv Complexity (A) and slope low band exponent (B), slope high band exponent (C), DTF outflow (D), alpha peak (E), delta (F), theta (G), alpha (H), beta (I) and gamma (J) power between awake-unparalysed, awake-paralysed and sedated-paralysed. The measures were visualised as a a spatial distribution on a scalp topography (left) and a distribution of the frequency of values for each condition (the median is indicated as a white dot, right). Statistical tests (paired t-test) were performed between conditions. Values were aggregated across all channels and epochs resulting in one value per participant and condition (6 values per condition). P-values were Bonferoni corrected for six comparisons (ns *p* > .05, * *p* ≤ .05, ** *p* ≤ .01, *** *p* ≤ .001, **** *p* ≤ .0001). For a measure to be considered successful, it should remain unchanged between awake-unparalysed (AU) and awake-paralysed states and only differ when comparing either awake state (AU/AP) to the sedated-paralysed (SP) state (e.g. Delta power). For a measure to be considered misleading AU needs to differ from AP and AP not differ from SP (e.g. LZc).

### 3.3 Prediction of participant state

We then predicted the state of the participants from 5 sec epochs using a SVC in a leave-one-participant out cross-validation approach. This adds the temporal and spatial dimension that was not considered in the statistical analysis.

#### 3.3.1 Naïve: Training and testing on awake-unparalysed and sedated-paralysed

Prediction accuracy was near to perfect for most measures with the exception of peak alpha frequency. See **Figure 4** 2 × 2 confusion matrices.

**Figure 4.**
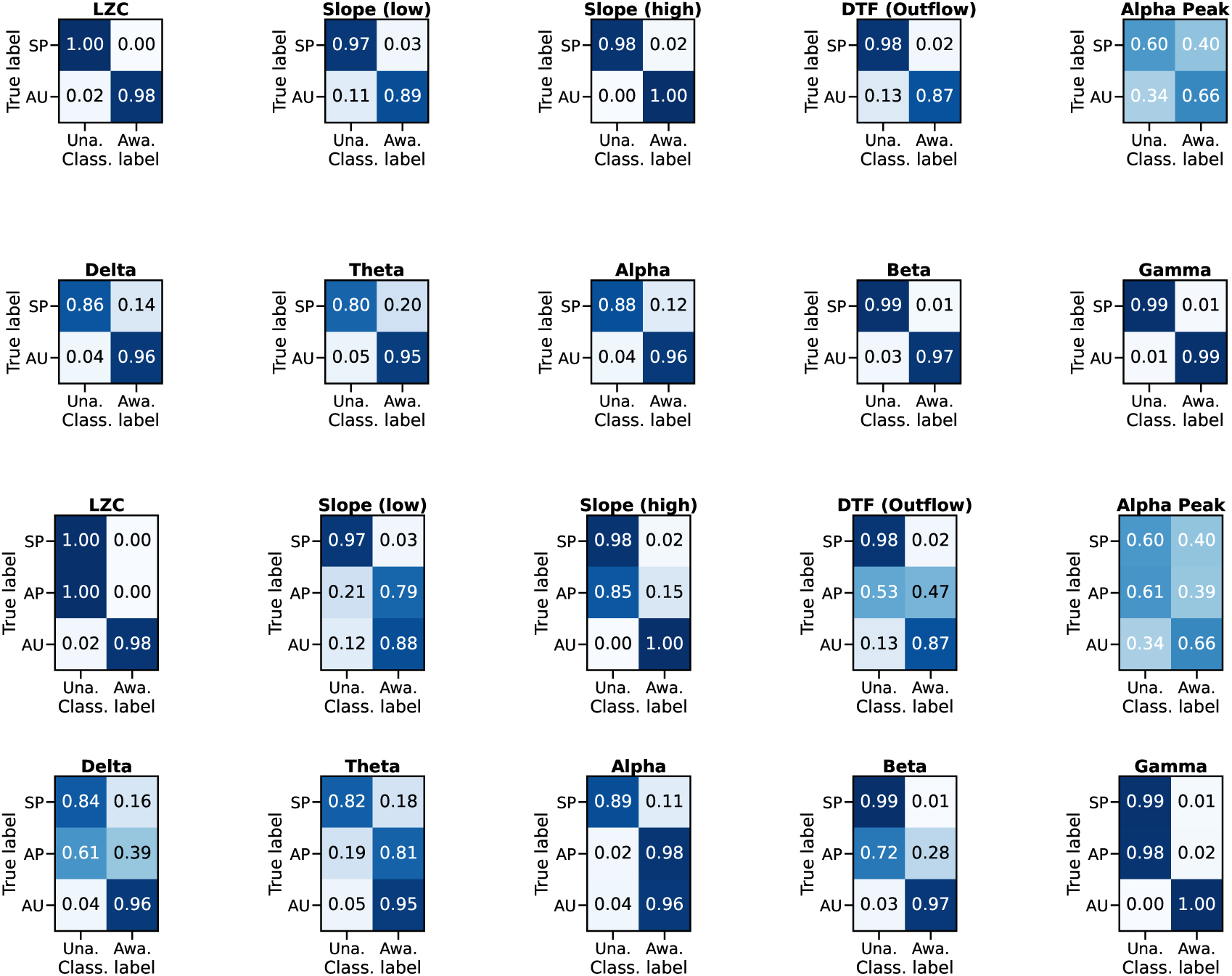
Confusion matrices that were computed by classifying individual epochs in a leave-one participant out cross-validation. The values reported in the matrix were computed as the mean across all cross-validation steps. The awake-paralysed (AP) state was excluded from training and testing (naïve approach, 2 × 2 matrices) or excluded from training and included in testing with the same label as the awake-unparalysed (AU) state (two class approach, 3 × 2 matrices). AP and AU trials predictions were rated as true when the prediction label was aware (awa.). Sedated-paralysed (SP) trials were rated as true if labelled as unaware (una.). Results were computed using the measures indicated in the title of each matrix. For a measure to be considered successful, it should classify both the awake-unparalysed and awake-paralysed states as aware, and the sedated-paralysed state as unaware. The naïve approach led to near perfect prediction using diversity, slope (high), beta and gamma power. Prediction using peak alpha frequency was poor. The two class approach revealed most measures predicted the awake-paralysed as unaware. The only measure to achieve high accuracy in this task was using alpha power.

#### 3.3.2 Two classes: Training on awake-unparalysed and sedated-paralysed and testing on all three states

The question we wanted to answer using this approach was whether a classifier trained on the awake-unparalysed and sedated-paralysed states would predict the awake-paralysed state as unaware or aware. Predicting awake-paralysed as aware would mean the measure might reliably detect AAGA, making this a successful measure according to our assessment. This means that only alpha power could reliably predict the state resembling AAGA in this dataset. See **Figure 4** 3 × 2 confusion matrices.

#### 3.3.3 Exploratory analysis of 55 features

The performance of each measure was quantified using the *F*_1_ score (see **Figure 5**) both for the naïve and the two class approach. The highest performance was achieved with FOOOF features averaged across the 1-20 Hz band. LZc and permutation entropy dropped considerably in the two class approach. See supplementary material for additional details.

**Figure 5.**
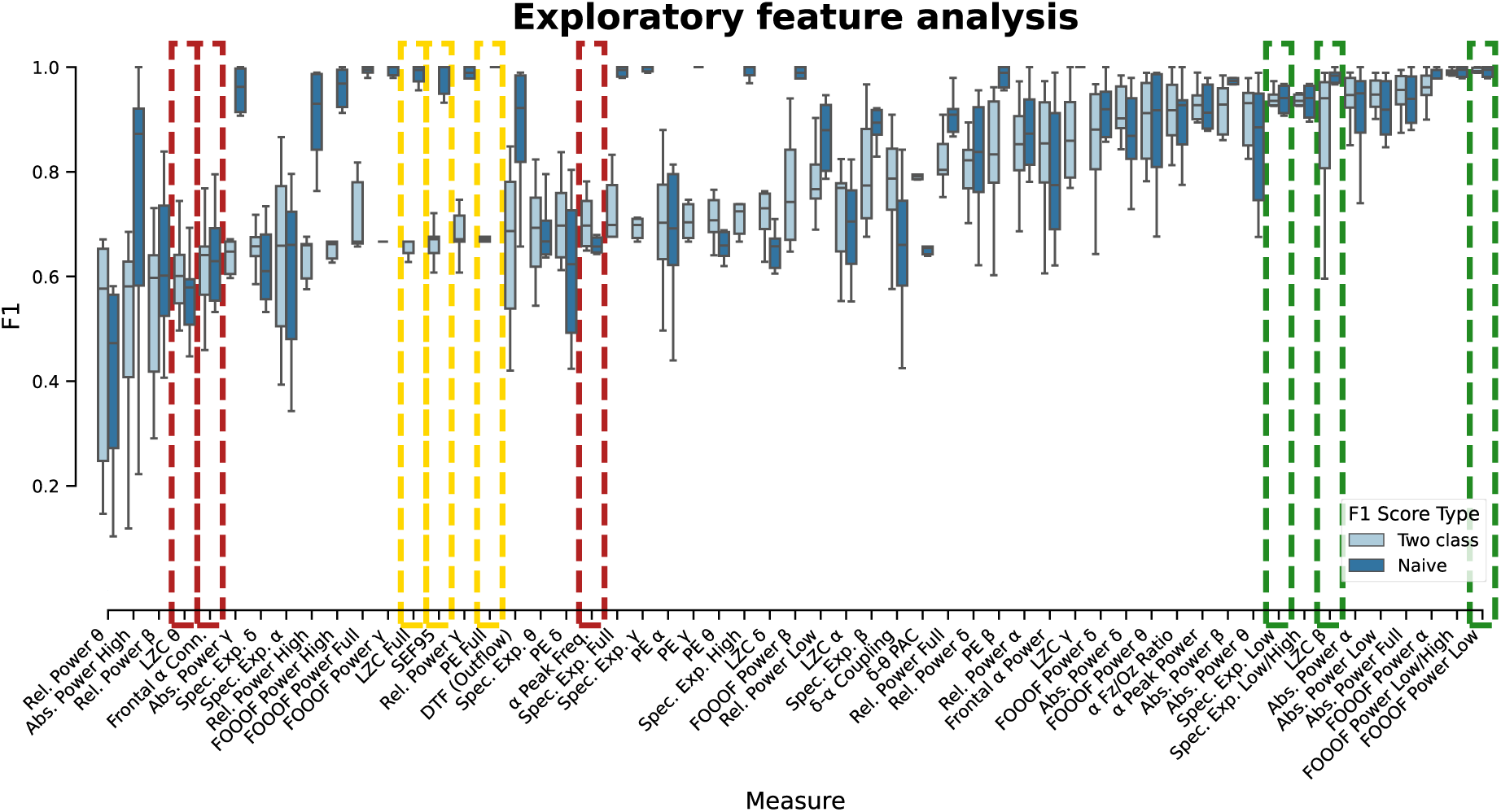
Exploratory feature analysis of classification performance. Box plots show the distribution of F1 scores for various electroencephalogram (EEG) derived measures used to classify states. F1 scores are differentiated according to training protocol: naive (dark blue, awake-paralysed (AP) state was excluded from training and testing) and two class (light blue, AP excluded from training and included in testing with the same label as the awake-unparalysed (AU) state; they can be considered as a measure of success in the context of our investigation and the measures are sorted from right to left according to success). We visualised the distributions of nine exemplary measures in the supplemental materials. These measures were highlighted with a coloured dashed box. Green: reliably predict awake-paralysed as awake, yellow: reliably predict sedated-paralysed as distinct from awake-unparalysed but confuse with awake-paralysed; red: confuse all three states.

## 4 Discussion

Our findings revealed that while many measures could readily distinguish awake-unparalysed from sedated-paralysed, they failed to recognise awareness in the awake state with paralysis, resembling AAGA in patients. Notably, alpha power was the most robust measure for predicting awareness in the paralysed wakefulness state with an exploratory analysis indicating that 1-20 Hz band power was the most successful. This work highlights the potential limitations of current EEG-based measures for detecting AAGA.

### 4.1 Why was awareness not detected in the awake-paralysed state?

EEG has been used to study effects of anaesthetic agents on brain activity for more than 85 years.^28,29^ However, it has proven difficult to find an EEG-based monitor of consciousness. One reason for this is probably EMG contamination of the EEG signals.^16^ This is the main cause of the unreliability of the widely used commercial indices **^schuller respons 2023 that depend on “cgls –emg” to detect awareness.^**^, 14^ We hoped that by focusing on slow (alpha band) oscillations, our putative DTF-based marker of consciousness should avoid serious EMG contamination.^20,30^ However, having now investigated this directly we find that it is indeed influenced by muscle activity, leading to misclassification of aware vs. unaware states. As another example, LZc changed significantly between awake-unparalysed and awake-paralysed. There was no significant difference between awake-paralysed and sedated-paralysed, which was then reflected in classification with the SVC in **Figure 4** 3 × 2 matrix. We observed the strongest decrease of the measure on peripheral electrodes, near major muscles, suggesting that the randomness measured with this method was also influenced by EMG.

### 4.2 Are our results meaningful despite small sample size?

While our study demonstrated the limitations of current EEG-based measures, such as LZc with high power, larger studies are needed to confirm generalisability and evaluate the influence of factors such as the choice of general anaesthetic agent.

### 4.3 Were our participants unconscious?

The propofol doses used in this study, particularly for participant 0, were more consistent with deep sedation than with surgical anaesthesia. This was intentional, as our aim was to achieve a level of sedation sufficient for tolerance of the LMA and maintenance of intermittent positive pressure ventilation until safe reversal of NMB could be accomplished in a non-operating theatre environment. While unconsciousness may occur at these concentrations in healthy volunteers^31,32^ we acknowledge the limitation that, due to the ischaemic limit of the IFT, we could not objectively verify unresponsiveness during the sedated-paralysed state. Although all 6 participants afterwards recalled no experience (including TOF testing) from the propofol sedation period, retrospective participant reports may be confounded by the amnestic effects of propofol. As such, our findings regarding the effects of propofol combined with NMB (sedated-paralysed) on EEG measures should be interpreted as reflecting deep sedation rather than definitive surgical anaesthesia.

### 4.4 What does this mean for monitoring patients?

The methods we investigated in this study were computed from the spontaneous EEG and can thus be utilised for time resolved monitoring. The implications for monitoring AAGA became apparent when we added the paralysis condition to the test set (**Figure 3 and 4** 3 × 2 matrices). Here we observed an increased bias towards categorising the awake-paralysed state as unaware, with the exception of features based on alpha power. This was consistent over all participants. The analysis of alpha log(power) in **Figure S3** supported the observed increase between awake-paralysed and sedated-paralysed states enabling the SVC to differentiate the two states which could be used to detect AAGA.

The anteriorisation of alpha induced by propofol is a commonly observed phenomenon.^33,34^ Unfortunately, decreases in alpha have also been observed.^35^ The impact on alpha has also been reported to be drug dependent: 4% of patients anaesthetised with sevoflurane or desflurane did not exhibit any alpha.^36^ The effect of Xenon on alpha appears to be inconsistent.^37,38^ Ketamine appears to cause changes in alpha peak frequency.^18^ In conclusion, it is unlikely that alpha log(power) is a reliable marker of awareness for all anaesthetic agents. .

## 5 Conclusions

We found, when the awake-paralysed state was included in the test set, nearly all measures failed to correctly classify it as aware. We can thus only conclude that EMG contamination of EEG has a surprisingly strong influence on EEG-based measures, and that the perfect measure of consciousness and marker of AAGA has not yet been found. Only log(power) features in the alpha band achieved high accuracies (**Figure 4**). Since different anaesthetics vary in their impact on the alpha rhythm, it is not reasonable to generalise this observation. In an exploratory analysis we found the average power in the 1-20 Hz band computed with FOOOF to have the highest accuracy at classifying the aware-paralysed state as aware (**5**) but this will also have to be verified with other anaesthetic agents. Our investigation highlights a critical limitation of relying on EMG-dependent indices, such as BIS, for the detection of AAGA as EMG activity is absent by design during NMB. Alternative EEG measures, potentially relying on tasks^39,40^ or perturbation,^41^ that are robust to the absence of EMG are needed for reliable monitoring of awareness.

## Supporting information

Supplemental Analysis

## Data Availability

All data produced in the present study are available upon reasonable request to the authors pending approval by the original authors.

## Authors’ contributions

Study conception (reanalysis): JFS

Study conception (data): JOW

Design of experiments: JOW

Administration of anaesthesia: AH

EEG data collection: KJP, JOW

Design of analysis: SH, BEJ

Data analysis: SH

Writing paper: SH, BEJ, AH, JOW, JFS

## Acknowledgements

The experiments were conducted in the Flinders University and Medical Centre, Adelaide, SA. Members of the team were: Whitham EM (neurologist), Fitzgibbon SP (cognitive scientist), Lewis T (computer scientist), Clark CR (psychologist), Lillie P (anaesthetist, decd), and Hardy A (anaesthetist). We thank Professor Johan Ræder, Oslo University Hospital, for helpful comments on the manuscript. For the purpose of open access, the author(s) has applied a Creative Commons Attribution (CC BY) licence to any Author Accepted Manuscript version arising.

