## Supplemental Analysis for "The effect of neuromuscular blockade on EEG-based measures of awareness"

### Appendix: Supplemental data

#### 1 Definition of important terms

Terms such as (un)consciousness, (un)awareness, and (un)responsiveness have previously been defined and used in various ways, sometimes causing conceptual confusion. Here, we will use the following definitions, which are in broad agreement with common language and much of the literature. "Conscious"/ "consciousness" is the basic state/ability to experience anything, including also dreaming and hallucinations, even when the subject is unable to sense or respond. "Awareness", however, involves the ability to experience at least some of the current reality, including sensations from the body such as pain, but excludes dreaming during deep sleep or anaesthesia. Thus, a deeply anaesthetised patient is "unaware" even when consciously experiencing a dream. "Responsiveness" is the ability to respond verbally or non-verbally to stimuli. Thus, a paralysed patient can be both conscious and aware while being unresponsive (connected vs disconnected-consciousness).[1] There are gray zones between these concepts, and further issues related to phenomenal consciousness vs. access consciousness,[2] reportability, and different types of memory, etc.,[3,4] but we will not address all these issues here.

The term spontaneous electroencephalogram (EEG) refers to activity recorded during rest, without any external stimulation or task.

#### 2 Investigation of artefact removal effects

##### 2.1 Pre-processing

We refer to the data processed to this point as 'Raw' in the context of the current analysis. In our attempts to reduce the impact of electromyogram (EMG) contamination[5] we investigated multiple approaches. (1) ICA: compute independent components using Infomax independent component analysis (ICA), see,[6] and label components using ICLabel[7] retaining only components labelled with 'brain' or 'other'. (2) ARICA: apply Autoreject automated artefact rejection,[8] repair only without rejection, apply ICA as in (1) but only on epochs not marked for rejection, run Autoreject again with same parameters. (3) EMG: compute independent components using Picard[9] and labelled using the `find_bads_muscle` function in MNE. The parameters of the labelling method were fine tuned using the EEG data in the current analysis, thus the approach may be considered as over-fitted for our analysis. And finally (4) CSDEMG: identical to (3) with the difference that we first transformed the data to current source density, see.[10]

#### 2.2 Impact of artefact removal

Removal of artefacts with ICA affected the awake-unparalysed state data more than the data from the awake-paralysed and sedated-paralysed state (see **Figure S1 A**). Bandpower decreased particularly in the peripheral electrodes in all canonical bands. The differences were small, also in the awake-unparalysed condition, and not significant in any band (see **Figure S1 B**). Overall, the difference in bandpower values in the gamma band between all three conditions decreased after artefact removal (see **Figure S1 C**).

In an explorative analysis we investigated further artefact removal methods and quantified their differences as changes in mean $F_1$ score at differentiating the aware (awake-unparalysed and awake-paralysed) and unaware (sedated-paralysed) states, see **Figure S2 (left)**. The line plot supports the the result of the statistical analysis as the differences in $F_1$ score between methods were quite small. None of the artefact removal methods were able to substantially affect the ability of the classifier to recognise the awake-paralysed state as aware. We investigated five feature types in more detail, see **Figure S2 (right)**. (1) The alpha peak frequency feature was not affected by the artefact removal but performance of this feature was generally low. (2) The slope feature performed nearly perfectly in the when the awake-paralysed state was excluded (naïve approach) but none of the artefact removal methods were able to alleviate the impact of including the awake-paralysed condition in the testing data. (3) Similarly, Lempel-Ziv complexity (LZc) features performed well in the naïve approach and did not improve after applying artefact removal. (4) Alpha bandpower features did not perform quite as well as (2) and (3) but performed the best after inclusion of paralysed-awake. (5) The outflow feature had the most variance in performance and was the feature to benefit the most from applying autoreject artefact removal.

#### 2.3 Why did the ICA based artefact removal only have a small effect?

Artefact removal affected mostly the awake-unparalysed state data on peripheral electrodes. The main issue that we observed in our analysis was the misclassification of awake-paralysed as sedated-paralysed, but the EEG recorded during these states was not as contaminated with muscle artefacts. Thus, the classification of the awake-paralysed state as aware using the features tested was not meaningfully improved by automatic muscle artefact removal from the EEG (see **Figure S2**).

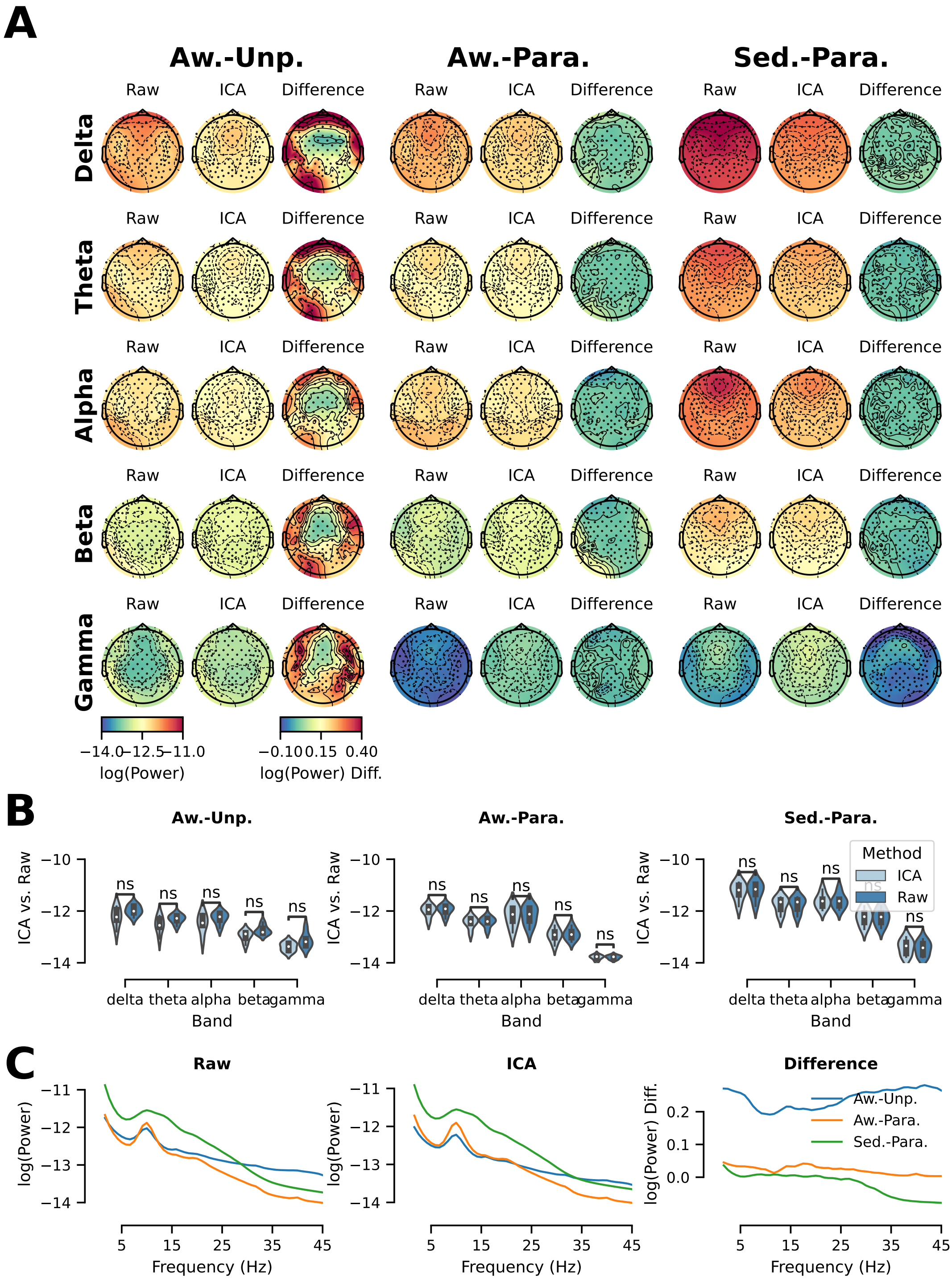


**Figure S1** – (A) Power spectral density (PSD) values were visualised as topographies for each canonical frequency band (rows), for each condition (awake-unparalysed, awake-paralysed and sedated-paralysed; columns) and within each condition before artefact removal with ICA (Raw), after artefact removal (ICA) and as the difference between both (Difference). (B) The distribution of the PSD values was visualised as a group of violin plots for each condition. Colours indicate whether the values of the raw (dark blue) or the ICA (light blue) cleaned data were displayed. Statistical tests (paired t-test) were performed between raw and ICA values. Values were aggregated across channels and epochs resulting in one value per participant and condition (six values per condition). P-values were Bonferoni corrected for 15 comparisons (ns $p > .05$, * $p \leq .05$, ** $p \leq .01$, *** $p \leq .001$, **** $p \leq .0001$) (C) PSD visualised as line plots and averaged across all epochs and channels. Raw, ICA artefact removal and difference values were visualised individually using separate lines per condition.

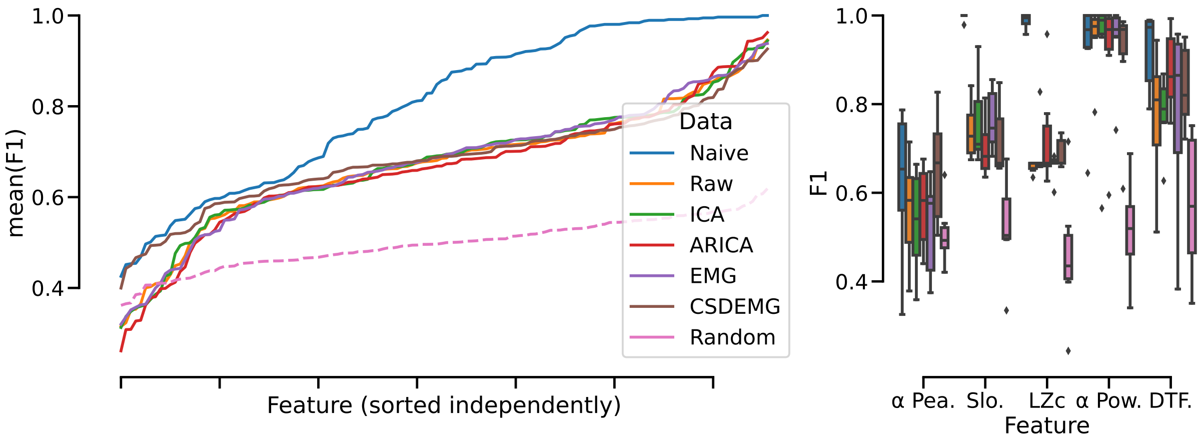


**Figure S2** – Compares mean $F_1$ score of naive approach (no awake-paralysed data in testing or training), raw data (awake-paralysed used in testing, no artefact removal) and data after artefacts were removed with IC Label (ICA), IC Label + autoreject (ARICA), MNE EMG detection (EMG), transform to CSD and then use MNE EMG detection (CSDEMG). $F_1$ scores were further computed with randomised training labels and the raw features (dashed pink line). Scores on all lines were sorted by mean, thus the x-axis may not always be for the same feature. This is of particular importance when interpreting the $F_1$ scores with randomised labels as one cannot conclude that they correlate with the non-randomised scores. On the right hand side, we show the exemplary impact on $F_1$ score in a box plot for alpha peak frequency, slope (low+high), LZc, alpha log(power) and DTF (outflow). Colours of boxes correspond to lines in left plot.

### 3 Individual differences alpha

We further explored the changes in alpha band power between conditions for each of the six participants individually, see **Figure S3**. For the statistical analysis we focused on the frontal region of interest (ROI). For all six participants, alpha power increased significantly between the awake-unparalysed and sedated-paralysed state. For three participants alpha power did not change, for two participants alpha increased and for one it decreased from awake-unparalysed to awake-paralysed state. For five of six participants alpha increased significantly from awake-paralysed to sedated-paralysed state.

For classification, the support vector classifier (SVC) leverages the spatial pattern of alpha activity, which typically showed anteriorisation (frontal $>$ posterior power) under propofol sedation. This pattern was dose-dependent: Participant 0, who received a smaller dose than the others, exhibited reduced anteriorisation (**Figure S3**). During leave-one-subject-out cross-validation, the classifier, trained on participants with stronger anteriorisation, struggled to distinguish P0's sedated-paralysed state from awake-paralysed. Consequently the $F_1$ score of participant 0 was the lowest with 0.8 (for reference P1: 0.95, P2: 0.99, P3: 0.99, P4: 1.00, P5: 0.95). This highlights a key limitation: alpha-based awareness detection assumes sufficient propofol dosing to induce the anteriorised alpha pattern characteristic of surgical anaesthesia. Future work should quantify dose-response relationships and explore whether spatial alpha patterns remain reliable at lower sedative doses.

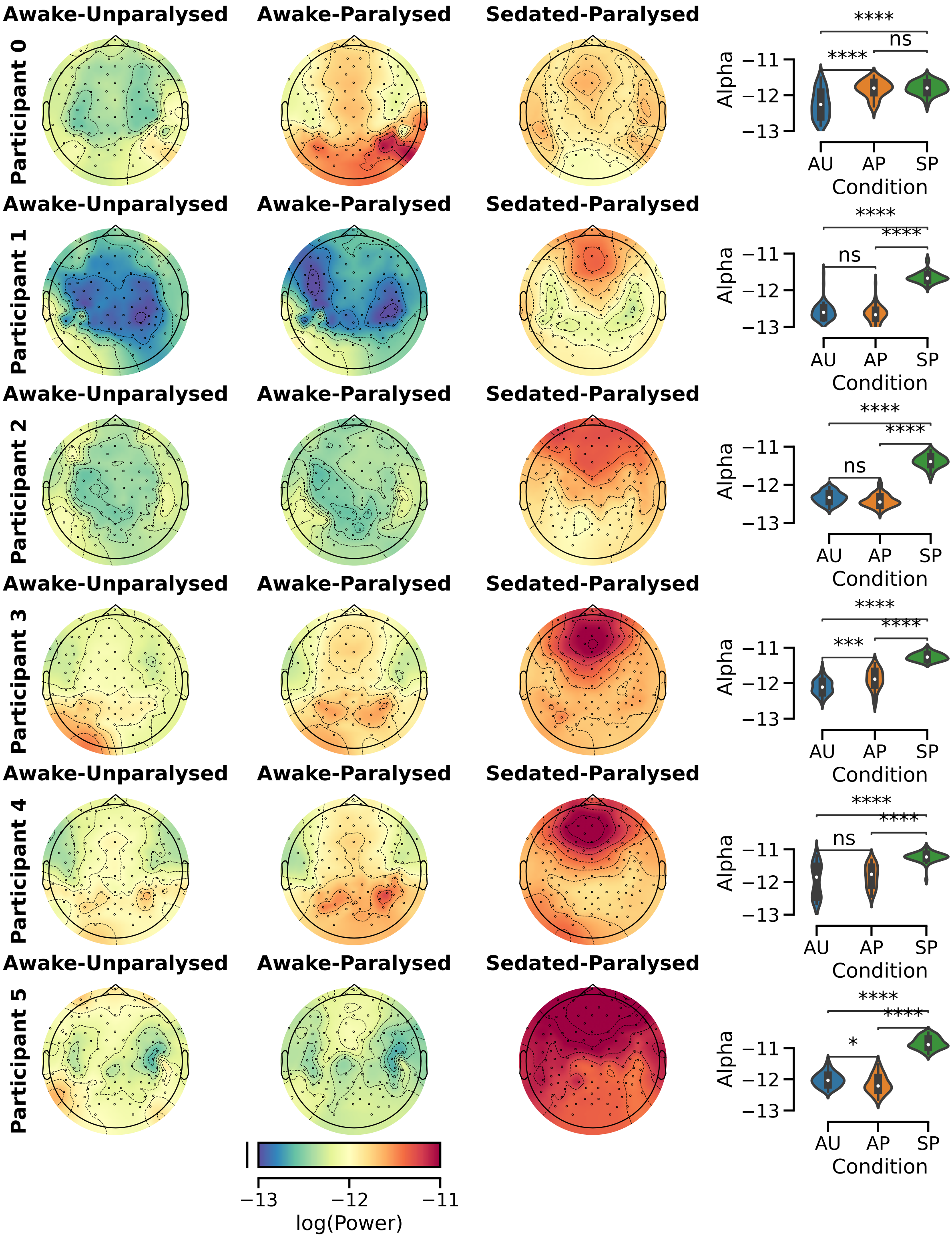


**Figure S3** – Power spectral density (PSD) values (as log band power computed using fitting oscillations & one-over-F (FOOOF)) in the alpha band visualised as three topographies, one per condition (awake-unparalysed (AU), awake-paralysed (AP), sedated-paralysed (SP), per participant. Numerical distributions of PSD values were visualised as violin plots again split according to condition. Statistical tests (paired t-test) were performed between conditions. Values were aggregated across channels (using only the frontal region-of-interest) resulting in one value per epoch and condition (47 values per condition). P-values were Bonferroni corrected for 18 comparisons (ns $p > .05$, * $p \leq .05$, ** $p \leq .01$, *** $p \leq .001$, **** $p \leq .0001$).

### 4 Three classes

In addition to the combinations of classes described in the main paper we also explored how well the classifier performed when we include awake-unparalysed, awake-paralysed and sedated-paralysed in training and testing with unique labels.

#### 4.1 Training and testing on all three states

Using this approach we investigated whether there were consistent differences in between the measures computed from each of the three states. One should note that it would be difficult to implement this approach on a larger sample as awake-paralysed data would need to be collected without anaesthesia/sedation. When inspecting the confusion matrices in **Figure S4** it is interesting to note which measures lead to confusion between sedated-paralysed and awake-paralysed (higher errors in the top left, measure less likely to detect accidental awareness during general anaesthesia (AAGA)) and which to confusion between awake-paralysed and awake-unparalysed (higher errors on the bottom right, measure more likely to detect AAGA). One can also regard only the errors when classifying the awake-paralysed state. If the error for predicting awake-paralysed as sedated-paralysed is higher this is more problematic for predicting AAGA than if the error is high for predicting awake-paralysed as awake-unparalysed. The former is the case for LZc, slope (high), directed transfer function (DTF) (outflow), delta, and gamma and the latter for slope (low), theta, alpha and beta. Again, predicting the state of the participant using alpha power led to the lowest error of awake-paralysed trials classified as sedated-paralysed (2%). The worst performance was again obtained using the alpha peak measure.

### 5 Exploratory analysis of ROIs

#### 5.1 ROIs

In addition to the features described in the main article, we performed an exploratory analysis of six features (alpha peak frequency, spectral slope low and high combined, FOOOF alpha power, FOOOF 1-45 Hz (full), LZc and DTF (outflow)) restricted to six ROIs. See **Figure S5** for the location of each ROI.

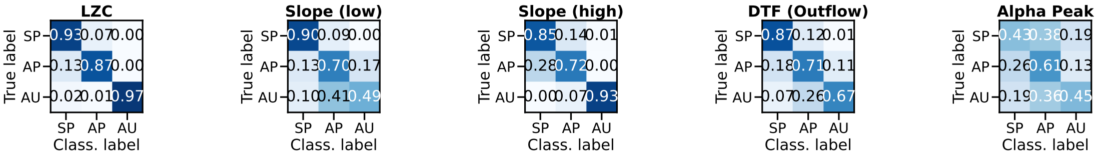


**Figure S4** – Confusion matrices that were computed by classifying individual epochs in a leave-one participant out cross-validation scheme. The values reported in the matrix (AU: Awake-Unparalysed, AP: Awake-Paralysed, SP: Sedated-Paralysed) were computed as the mean across all cross-validation steps. All three states were included in training and testing. Please note that this training and testing approach could not be implemented with a larger sample as it is unlikely that the awake-paralysed state will be available. Results were computed using the measures indicated in the title of each matrix.

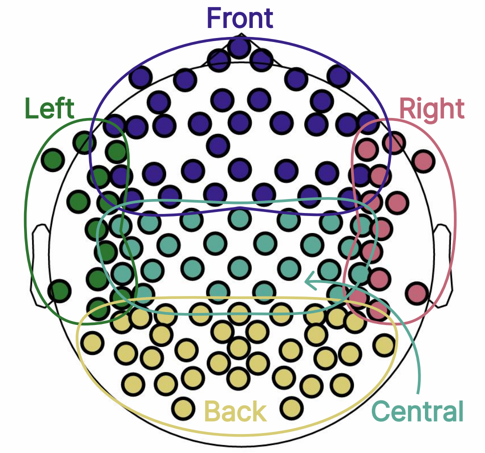


**Figure S5** – Regions-of-interest (ROIs) as used in the exploratory analysis. Electrodes were assigned to frontal (blue), left (green), central (turquoise), right (pink) or occipital (yellow) ROIs.

##### 5.2 Results

When exploring the other feature types using only sedated-paralysed and awake-unparalysed for training and testing (see **Figure S6**, top) we obtained a median $F_1$-score of 0.93 (SD 0.11, range 0.56 to 1.0) for alpha power, 1.0 (SD 0.01, range 0.95 to 1.0) for 1-45 Hz power, 1.0 (SD 0.02, range 0.91 to 1.0) for slope, 0.55 (SD 0.19, range 0.13 to 0.88) for alpha peak frequency, 0.99 (SD 0.04, range 0.84 to 1.0) for LZc and 0.87 (SD 0.13, range 0.46 to 0.99) for DTF (outflow). Across all feature types the highest median $F_1$-scores were obtained using frontal and full ROIs (0.98) and lowest using occipital ROIs (0.9). Inclusion of awake-paralysed epochs in the test set lead to reduced $F_1$-scores (see **Figure S6**, bottom): 0.93 (SD 0.09, range 0.66 to 1.0) for alpha power, 0.75 (SD 0.12, range 0.65 to 1.0) for 1-45 Hz power, 0.74 (SD 0.08, range 0.66 to 0.96) for slope, 0.43 (SD 0.17, range 0.07 to 0.71) for alpha peak frequency, 0.67 (SD 0.08, range 0.54 to 0.93) for LZc and 0.68 (SD 0.05, range 0.52 to 0.78) for DTF outflow. The range of $F_1$ scores acros ROIs was quite narrow (0.67-0.73). For most measures frontal or full-head was the best choice.

#### 6 Classification

Classification was performed using scikit-learn.[11] We applied a standard scaler, used a linear SVC for feature selection (L1 penalty), and a linear SVC for classification (C=1, gamma='auto').

We evaluated the accuracy of classification using confusion matrices and $F_1$ scores. The $F_1$ score was computed as the harmonic mean of precision and recall.[12] In the context of awareness detection we can view precision as the number of time segments in which a patient is aware that were classified as aware divided by the total number of time segments that were classified as aware. A high precision value means the classifier is mostly correct when predicting awareness but may miss times of awareness. Similarly we can view recall as number of aware time segments classified as aware divided by the total number of aware time segments. A high recall value means the classifier will not miss segments of awareness but may mistake segments of unawareness as aware. Since the $F_1$ score combines both metrics a high $F_1$ score means the classifier does not miss segments of awareness and does not mistake segments of unawareness for aware.

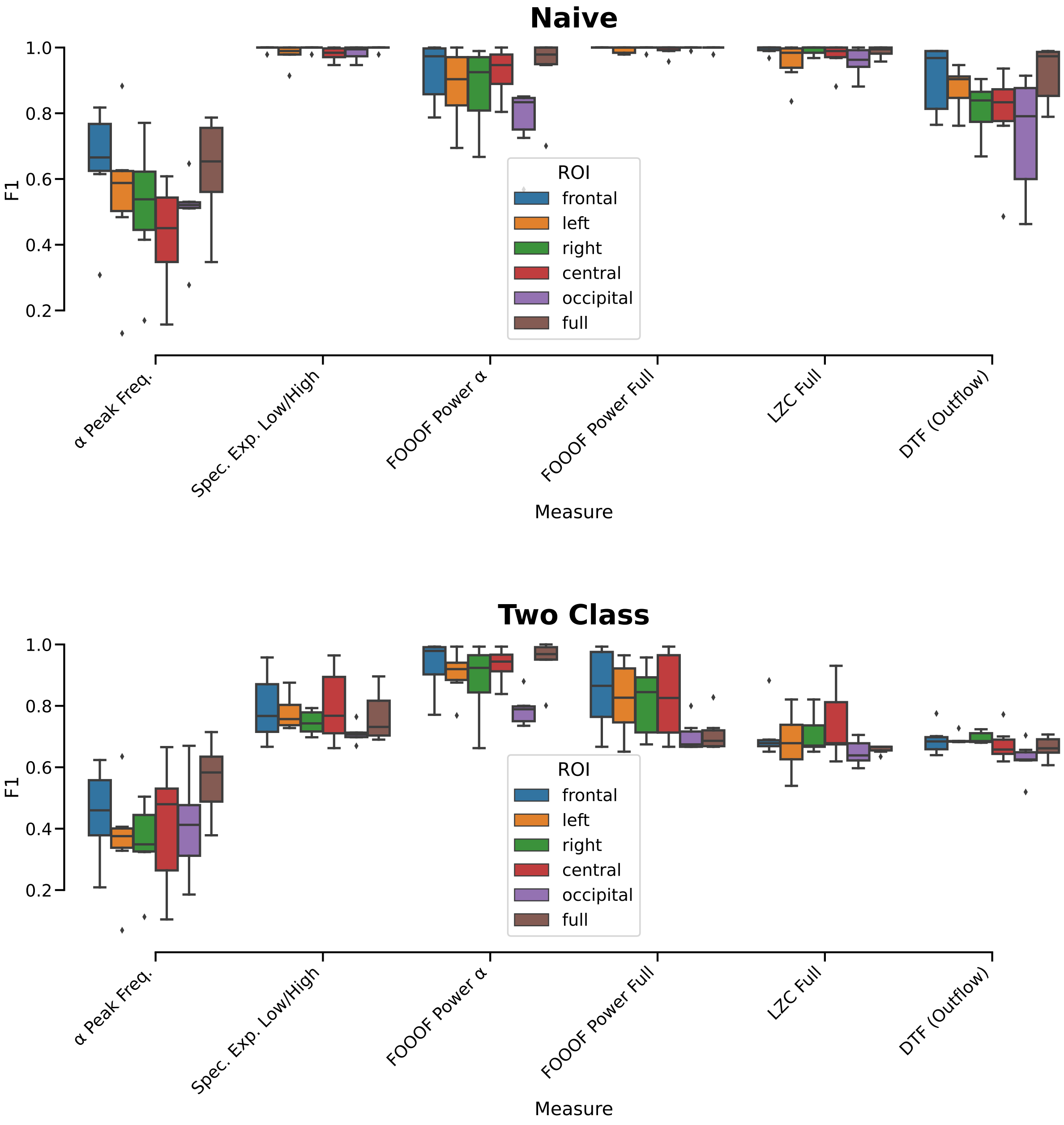


**Figure S6** – $F_1$ scores of classification of individual epochs in a leave-one participant out scheme using a six features computed using six different ROIs (**Figure S5**). The $F_1$ scores were computed by either training and testing the classifier on awake-unparalysed and sedated-paralysed epochs (naïve, top) or on awake-unparalysed and sedated-paralysed epochs and tested on awake-unparalysed, awake-paralysed and sedated-paralysed epochs (two class, bottom). Paralysis-awake trials predictions were rated as true when the prediction label was aware. The bars were coloured according to ROI.

### 7 Features

#### 7.1 Main feature details

We extracted five sets of features. (1) Logarithmic *bandpower* from the canonical frequency bands (delta 1-3.5 Hz, theta 3.5-8 Hz, alpha 8-13 Hz, beta 13-26 Hz, gamma 26-45 Hz) and the full band from 1-45 Hz. Power spectral density (PSD) was computed for each channel and epoch using the Welch method (resulting in 53 frequency bins). We then used FOOOF[13] to parameterise the power spectra for each epoch and channel and stored the logarithmic bandpower for each frequency bin. (2) *Spectral slope* was computed using identical processing with the difference that we stored the exponent of the aperiodic component for each channel and epoch for each of the canonical bands, the full band (1-45 Hz), the low half (1-20 Hz) and the high half (20-40 Hz)[14] using Python scripts kindly provided by the authors to compute the (negative) spectral exponent $\beta$. (3) *Peak frequency* was again computed using identical processing as (1) with the difference that we stored the frequency of the peak of each of the bands in Hz as determined with FOOOF. (4) *Signal diversity* was computed for each epoch and channel on the time domain signal using single channel LZc[15] with Python scripts kindly provided by the original authors. (5) *DTF (Outflow)*,[16] was computed by estimating brain connectivity with DTF[17] and computing the median of all outgoing connections for each electrode. We computed DTF between 8-12 Hz and re-referenced the data to FC1, for a subset of 19 EEG channels making the procedure as similar as possible to the original work, using the Python script provided by the author.

#### 7.2 Group statistics

Please refer to **Tables S1** and **S2** for group level values and statistics.

#### 7.3 Exploratory analysis of features

Inspired by the selection of measures presented by Casey et al., 2024[19] we expanded our analysis to an exploration of a total of 55 features and compared their performance using the $F_1$ score computed by either training and testing the classifier on awake-unparalysed and sedated-paralysed epochs (naïve approach) or on awake-unparalysed and sedated-paralysed epochs and tested on awake-unparalysed, awake-paralysed and sedated-paralysed epochs (two class approach). All measures were computed scripts implemented in Python with the MNE toolbox.[20] Additional modules were used where indicated. *SEF95*: We computed the PSD using Welch's method on the 1 to 45 Hz band.[21,22,23] We note that other

**Table S1** – Measures of awareness in three conditions: (1) awake-unparalysed, (2) awake-paralysed, and (3) sedated-paralysed. Median values were computed across all epochs and electrodes for each participant which were then used to compute median, standard-deviation (SD) and range from minimum to maximum in the table below.

| **Measure** | **Statistic** | **awake-unparalysed** | **awake-paralysed** | **sedated-paralysed** |
|---|---|---|---|---|
| LZc | Median | 0.84 | 0.62 | 0.55 |
| | SD | 0.12 | 0.08 | 0.06 |
| | Range | 0.33 to 0.99 | 0.32 to 0.83 | 0.32 to 0.77 |
| Spectral Slope (Low) | Median | -1.14 | -0.95 | -1.08 |
| | SD | 0.56 | 0.44 | 0.53 |
| | Range | -4.67 to 0.83 | -4.01 to 0.83 | -3.13 to 0.89 |
| Spectral Slope (High) | Median | -0.55 | -4.02 | -5.2 |
| | SD | 1.38 | 1.24 | 1.09 |
| | Range | -6.69 to 3.32 | -9.41 to 0.84 | -10.39 to 3.61 |
| Median Outflow | Median | -0.93 | -0.79 | -0.72 |
| | SD | 0.35 | 0.23 | 0.23 |
| | Range | -2.25 to -0.11 | -2.14 to -0.05 | -2.22 to 0.0 |
| Alpha Peak | Median | 10.14 | 10.15 | 10.48 |
| | SD | 1.13 | 1.05 | 1.3 |
| | Range | 8 to 13 | 8.0 to 13.0 | 8 to 13 |
| Delta Power | Median | -11.96 | -11.95 | -11.19 |
| | SD | 0.44 | 0.37 | 0.49 |
| | Range | -13.12 to -8.31 | -13.06 to -8.98 | -12.7 to -6.79 |
| Theta Power | Median | -12.3 | -12.4 | -11.73 |
| | SD | 0.34 | 0.32 | 0.38 |
| | Range | -13.3 to -8.93 | -13.4 to -9.81 | -12.8 to 7.71 |
| Alpha Power | Median | -12.67 | -12.2 | -11.63 |
| | SD | 0.5 | 0.58 | 0.43 |
| | Range | -13.37 to -9.03 | -13.61 to -9.73 | -13.02 to -8.39 |
| Beta Power | Median | -12.75 | -12.92 | -12.25 |
| | SD | 0.35 | 0.39 | 0.52 |
| | Range | -13.73 to -8.9 | -13.99 to -10.81 | -13.93 to -8.65 |
| Gamma Power | Median | -13.15 | -13.79 | -13.43 |
| | SD | 0.43 | 0.32 | 0.47 |
| | Range | -14.14 to -9.12 | -14.52 to -11.35 | -14.69 to -8.78 |

studies used 1-55 Hz but this overlaps with line noise in many countries. We computed the cumlative trapezoid (using Scipy) over the PSD, normalised each value by the total, and determined the frequency at which the cumulative power crosses the 95th percentile. The log transformed values were used for classification with one value per channel (all channels) per epoch.

*Absolute and relative band power:.* As an alternative to our approach using FOOOF we also computed absolute and relative band power in all canonical frequency bands.[24,25,26] We computed the PSD using Welch's method on the 1-45 Hz band and extracted the mean for each frequency band and the mean of each band divided by the total power across all bands. This resulted in one value of absolute and relative power per canonical band, channel (all channels used) and epochs.

*Alpha peak power and frequency:* To align with the approach of previously published work we used Welch's method to compute the power in the Alpha band. We used numpy to compute the maximum value per epoch and channel and stored peak power and frequency. We further computed the log ratio of Fz and Oz.

*Frontal Alpha power:* Computed the mean power in the alpha band over eight frontal channels (AFp1, AFp2, Fz, FC5, FC3, FCz, FC4, FC6) resulting in one value per epoch.

*Spectral slope/exponent:* See description in main paper. We used the script from Colombo et al.[14] to compute one value per band, channel (all) and epoch. As proposed by Colombo et al. also computed the slope for the low (1-20 Hz) band and the high (20-40 Hz) band and also combined both in one feature set. This then either resulted in one or two values per band, channel (all channels) and epochs.

*FOOOF:* This was computed as described in the main paper with the difference that to align the number of features with the other methods in this comparison we averaged the frequency bins of each band.[13] We further computed the mean of the low and high band and combined both in one feature set. This then either resulted in one or two values per band, channel (all channels) and epochs.

*LZc:* We computed LZc[15,27] the scripts provided by Schartner et al. as described in the main paper with the addition of computing the complexity with the epochs band pass filtered to the canonical bands. This resulted in one value per channel (all) and epoch.

*Permutation Entropy:* We computed permutation entropy[28,29] using the AntroPy toolbox maintained by Raphael Vallat. We computed one value per band, channel (all) and epoch.

To compute the connectivity metrics we either used a set of eight frontal channels (AFp1, AFp2, Fz, FC5, FC3, FCz, FC4, FC6), eight occipital channels (P1, Pz, P2, PO7, PO8, O1, Oz, O2) or a distributed subset (F7, F3, Fz, F4, F8, T7, C3, Cz, C4, T8, P9, P7, P3, Pz, P4, P8, P10, O1, O2).

*Frontal Alpha connectivity:* Computed weighted phase lag index[30] in the alpha band between all pairs in the frontal channel set and averaged resulting in one value per epoch. *Delta-Alpha peak Coupling:* We filtered the epochs to the delta and alpha band, computed the Hilbert envelope of the alpha epochs and computed the correlation with the delta epochs.[31]

*Delta-Theta phase-amplitude coupling:* We filtered the epochs to the high delta (2-4 Hz) and theta band, used Hilbert transformation to compute the phase of the delta epochs and the envelope of the theta epochs, assigned the phases to 18 bins, normalised each bin with the sum across all bins, computed the sum of each bin multiplied with the log of each bin and normalised with the one over the log of the number of bins.[31]

*DTF (Outflow):* Computed as described in main paper[16] on distributed subset resulting in one value per channel per epoch.

Casey et al., 2024 further investigated global and local efficiency[32] and normalised symbolic transfer entropy.[33] We decided to omit both these measures as they require optimisation of parameters on the dataset. This is not possible when attempting to detect AAGA online. The parameters could be optimised on a separate subset of the data but we concluded this is outside the scope of our investigation.

##### 7.4 Results

The results were included in the main paper (**Figure 5**). Distributions of two exemplary measures (**Figure S7**) how changes in the distribution from awake-unparalysed to awake-paralysed caused decreased $F_1$ scores.

#### 8 Raw EEG traces

Exemplary five second EEG traces of 11 channels were extracted from each condition at 25%, 50% and 75% of the duration of the condition (**Figures S8-S13**).

#### 9 Are neuromuscular blocking agents (NMBAs) commonly used in clinical scenarios?

NMBAs have been administered to ease intubation and optimize patient positioning since the 1940s.[34] Use of NMBAs in intensive care units is regarded as controversial and there are recent publications arguing against their use in patients with critical illnesses.[35] In a study of patient satisfaction after anaesthesia and surgery 62% of n=10811 patients received NMBAs and 0.11% reported AAGA.[36] In another report of n=11440 clinical procedures n=7753 received NMBAs to assist with intubation.[37] In this sample 0.18% of patients that received NMBAs reported AAGA and 0.10% of patients that did not receive NMBAs. The authors further noted that recollection of awareness may change with the amount of time that passed since the surgery suggesting that the reported numbers constitute a lower bound of the incidence.

This is supported by observations that responses recorded using the isolated forearm technique (IFT) do

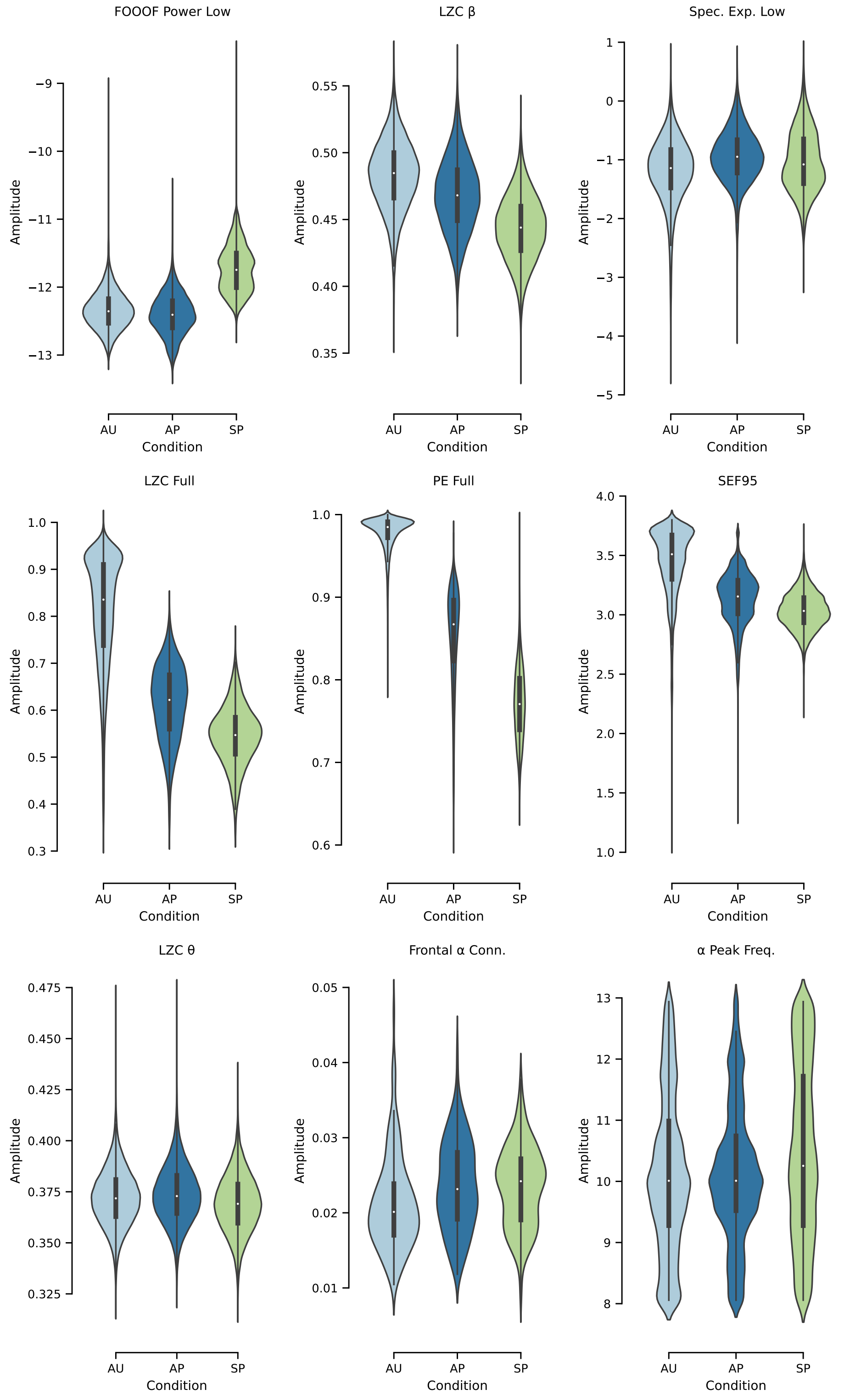


**Figure S7** – Nine measures selected to further explain the differences in performance. The distribution (including all participants, epochs and channels) of the measures the top row, changed less from awake-unparalysed (AU) to awake-paralysed (AP) than to the sedated-paralysed (SP) state. These were highlighted in green in **Figure 5**. The spectral slope of the low band changed mostly in spatial distribution (see **Figure 3** in main paper). The measures in the middle row changed between all three states which made it difficult to determine whether the participant was in the AP or the SP state, which decreased the $F_1$ score in our analysis and making it less probable that AAGA will be detected under the influence of neuromuscular blockade in clinical practice. These were highlighted in yellow in **Figure 5**. The measures in the bottom row did not reliably predict state changes due to the overlap of the distributions. These were highlighted in red in **Figure 5**.

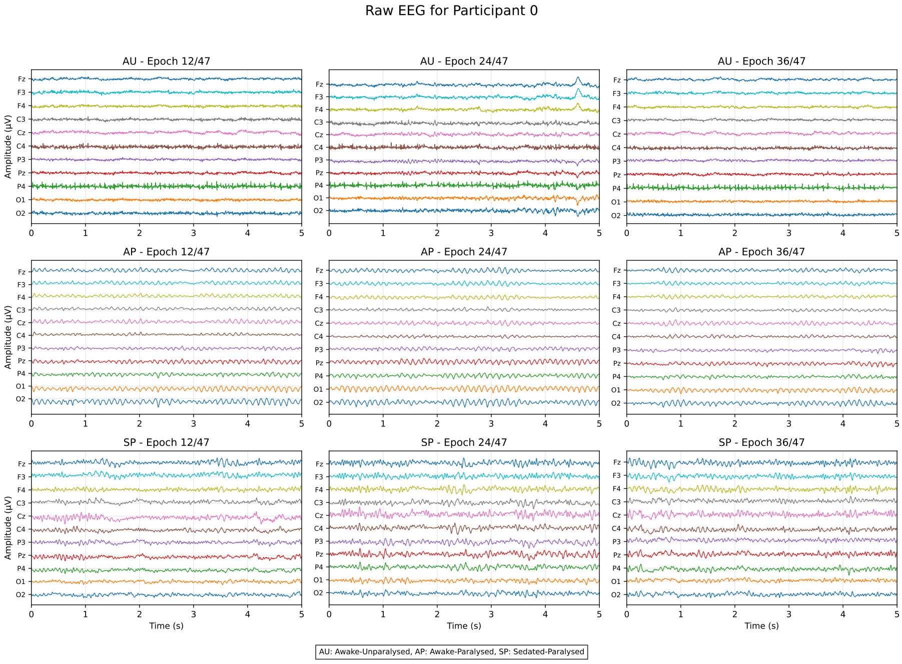


**Figure S8** – Raw electroencephalogram (EEG) traces from Participant 0. Representative 5-second epochs illustrate EEG patterns across 11 channels during Awake-Unparalysed (AU), Awake-Paralysed (AP), and Sedated-Paralysed (SP) conditions. Amplitude is shown in microvolts (μV) against time in seconds (s).

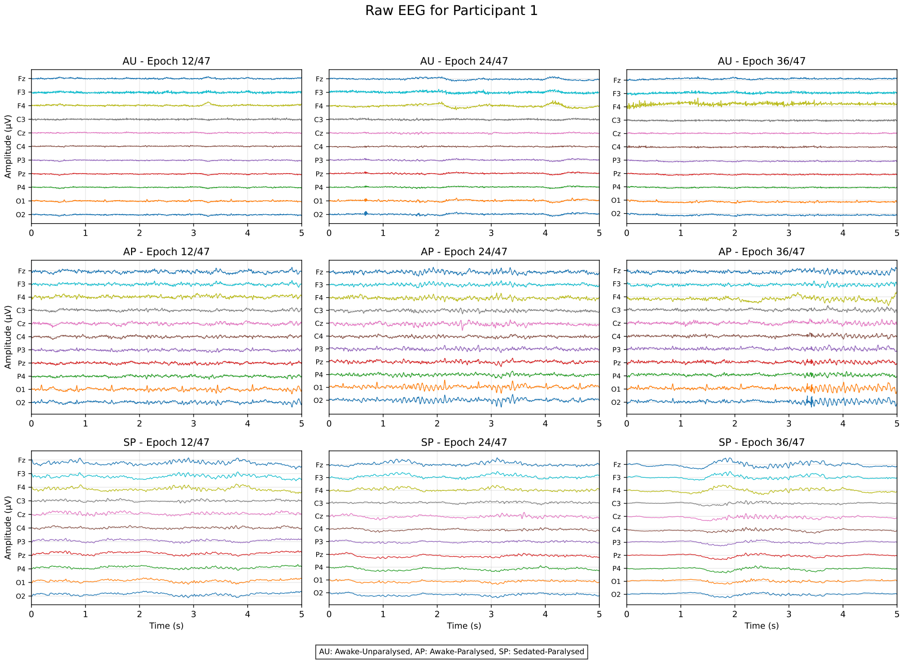


**Figure S9** – Raw electroencephalogram (EEG) traces from Participant 1. Representative 5-second epochs illustrate EEG patterns across 11 channels during Awake-Unparalysed (AU), Awake-Paralysed (AP), and Sedated-Paralysed (SP) conditions. Amplitude is shown in microvolts (μV) against time in seconds (s).

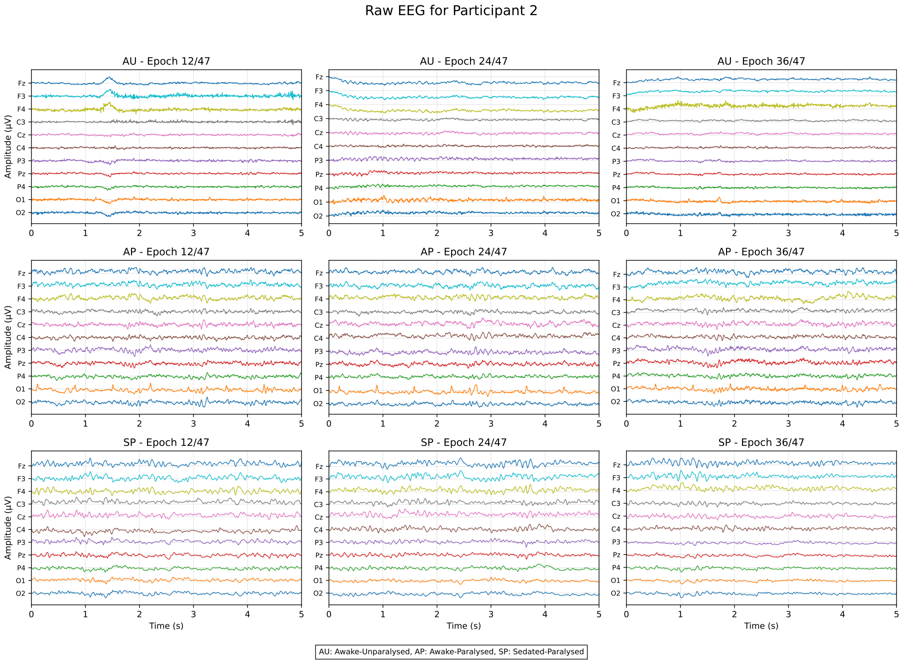


**Figure S10** – Raw electroencephalogram (EEG) traces from Participant 2. Representative 5-second epochs illustrate EEG patterns across 11 channels during Awake-Unparalysed (AU), Awake-Paralysed (AP), and Sedated-Paralysed (SP) conditions. Amplitude is shown in microvolts (µV) against time in seconds (s).

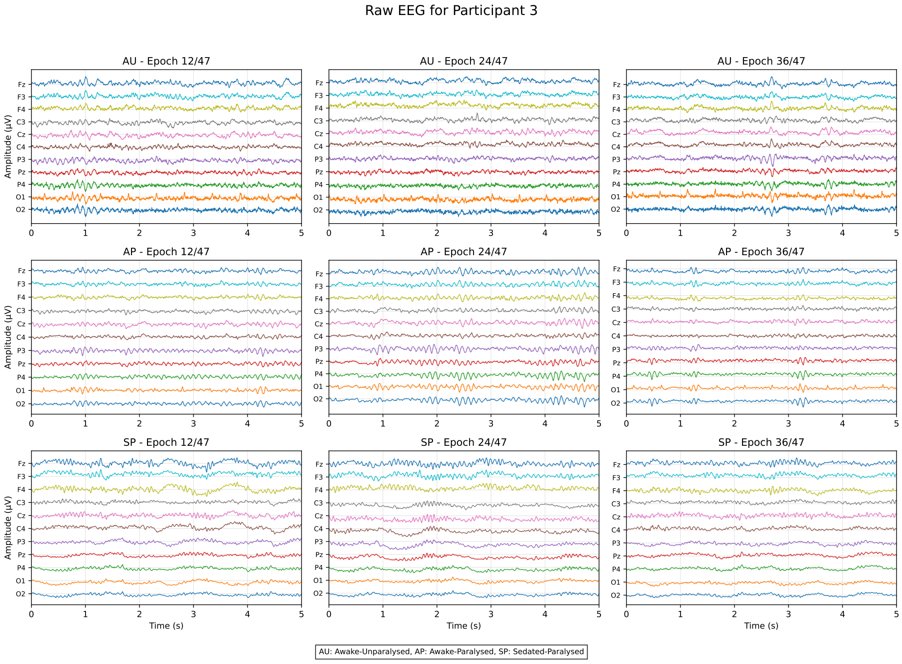


**Figure S11** – Raw electroencephalogram (EEG) traces from Participant 3. Representative 5-second epochs illustrate EEG patterns across 11 channels during Awake-Unparalysed (AU), Awake-Paralysed (AP), and Sedated-Paralysed (SP) conditions. Amplitude is shown in microvolts (µV) against time in seconds (s).

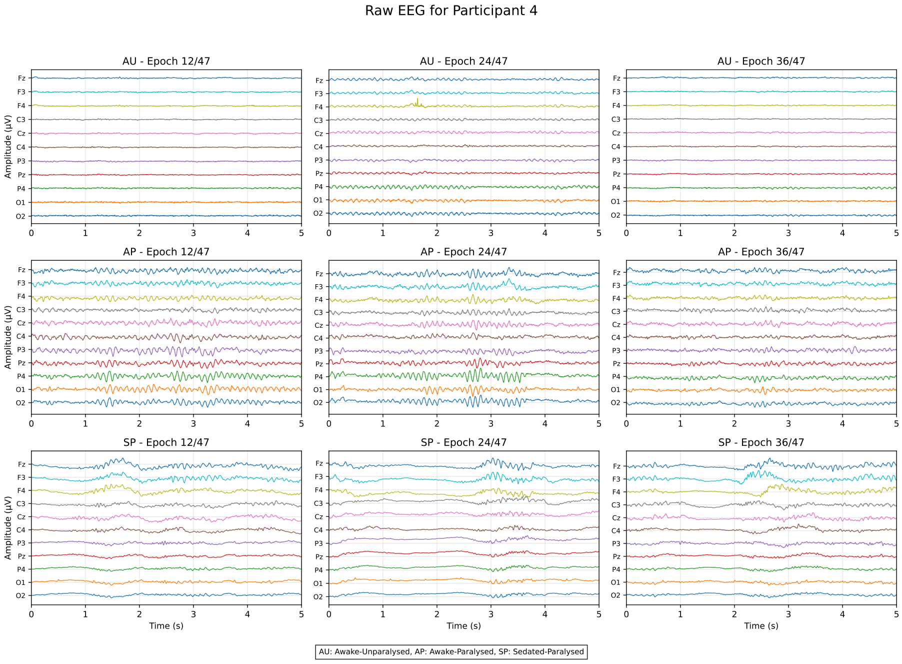


**Figure S12** – Raw electroencephalogram (EEG) traces from Participant 4. Representative 5-second epochs illustrate EEG patterns across 11 channels during Awake-Unparalysed (AU), Awake-Paralysed (AP), and Sedated-Paralysed (SP) conditions. Amplitude is shown in microvolts (µV) against time in seconds (s).

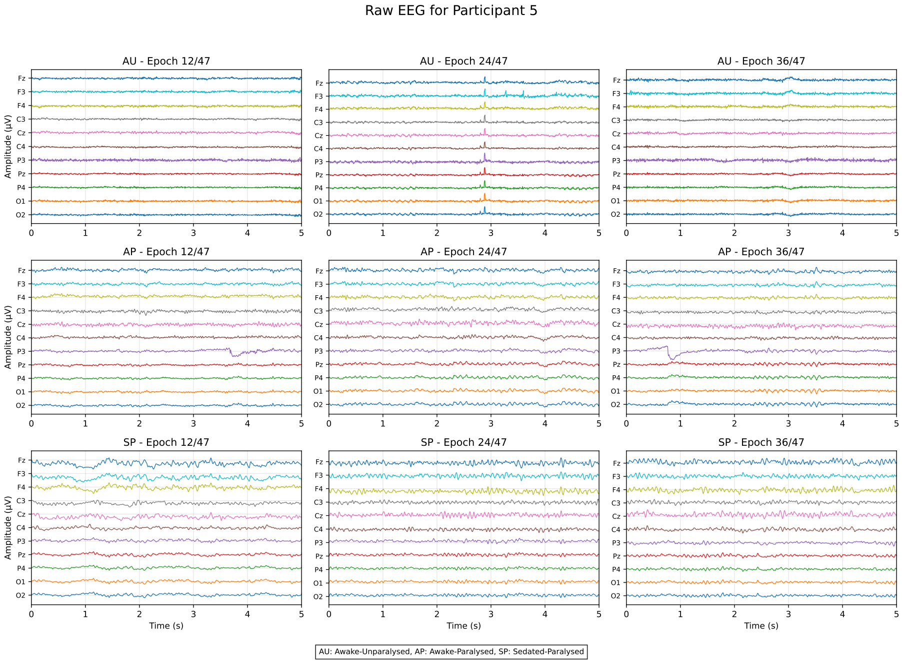


**Figure S13** – Raw electroencephalogram (EEG) traces from Participant 5. Representative 5-second epochs illustrate EEG patterns across 11 channels during Awake-Unparalysed (AU), Awake-Paralysed (AP), and Sedated-Paralysed (SP) conditions. Amplitude is shown in microvolts (µV) against time in seconds (s).

not coincide with recall or monitoring using bispectral index.[38] In regard to our study we conclude that NMBAs are frequently administered and constitute a risk factor for AAGA.

#### 10 Was the NMBAs dose similar to clinical scenarios?

Participants in our data set received a bolus dose of cisatracurium, 20 mg. This NMBA is also administered during surgery and to patients with critical illnesses to improve intubation conditions.[39,40] The dosage used during surgery can be lower than in our experiment, e.g. 0.1 mg.kg$^{-1}$ in,[41] which may affect how strongly measures of awareness are impacted by paralysis. The specific NMBA used may also have an impact on how the EEG is affected. NMBAs are generally categorised as either non-depolarizing (compete with acetylcholine at the receptor binding sites) or depolarizing (cause sustained depolarisation preventing further action potentials) blocking agents with the depolarizing agents leading to muscle fasciulations before paralysis.[42] Cisatracurium is categorised as a non-depolarizing agent and thus such fasciculations will not have impacted our recordings. Depolarizing agents are also less common, e.g. Suxamethonium is the only such agent available in the UK.[34] In summary, the NMBA administered to our participants is also utilised in clinical settings and is in fact representative for the most commonly used group of non-depolarizing agents. Thus, if a dosage resulting in a similar degree of twitch suppression is administered the effect on the EEG recording we observed should be representative for the use of NMBAs in clinical scenarios.

#### 11 Comparison with non-spontaneous EEG-based measures of consciousness

In addition to the use of spontaneous EEG, consciousness has been assessed by perturbational or task EEG-based measures.

The perturbational complexity index (PCI), developed by M. Massimini and colleagues[43] has been thoroughly tested and seems highly reliable for distinguishing conscious from unconscious brain states, including wake vs. sleep and general anaesthesia, and is probably less influenced by EMG.[44] However, because PCI requires multiple ($\sim$100) repeated stimuli (usually by transcranial magnetic stimulation) and averaging of EEG responses, it has a low time resolution, and is thus not so suitable for monitoring patients to rapidly detect AAGA.

Responses to sensory input and event-related potentials (ERPs) can also indicate altered awareness.[45,46]

Like PCI, these approaches require multiple trials, thus reducing time resolution.

Other approaches rely on active tasks, e.g. IFT, which does not require EEG or stimulation; but this can be used only for $\sim$20 minutes due to the need for reperfusion of the ischaemic isolated limb.[47] Also so-called brain-computer interfaces can be used to by-pass the need for muscular control, e.g. using motor imagery.[48] Investigations of dosage and anaesthetic agents may be needed to understand the mechanisms of disconnection and find the optimal way to detect AAGA.[49] In summary, alternative methods such as PCI and ERP[50] are proabably less affected by EMG, but suffer from low temporal resolution, more complex setup, or limited duration (IFT).

**Table S2** – Statistical comparisons between measures in the three different conditions: (1) awake-unparalysed (AU), (2) sedated-paralysed (SP) and (3) awake-paralysed (AP). Median values were computed across all epochs and electrodes for each participant which were then used to determine significance using Bonferoni corrected paired t-tests. Significant p-values are marked with a star (*). Cohen's $d$ and statistical power were computed with median and SD from **Table S1** using G*Power 3.1.[18] For a measure to be considered successful, it should remain unchanged between awake-unparalysed and awake-paralysed states (AU vs AP), and differ when comparing either awake-paralysed to the sedated-paralysed (AP vs SP). This criterion is highlighted in the statistical comparisons below. $^{-}$Measures with high power for AU vs AP shaded in red. $^{+}$ Measures with high power for distinguishing AP vs SP shaded in green.

| **Measure** | **Statistic** | **AU vs SP** | **AU vs AP** | **AP vs SP** |
|---|---|---|---|---|
| *LZc*$^{-}$ | $t_5$ | 10.08 | 6.62 | 2.12 |
| | $p$ | .001* | .007* | .526 |
| | $d$ | 2.8 | 2.1 | 0.9 |
| | $1-\beta$ | .99 | .98 | .48 |
| Spectral Slope (Low) | $t_5$ | −0.98 | −2.85 | 0.57 |
| | $p$ | 1 | .214 | 1 |
| | $d$ | 0.1 | 0.4 | 0.2 |
| | $1-\beta$ | .05 | .12 | .07 |
| *Spectral Slope (High)*$^{-}$ | $t_5$ | 10.39 | −4.73 | 5.18 |
| | $p$ | .001* | .031* | .021* |
| | $d$ | 3.7 | 2.6 | 1.0 |
| | $1-\beta$ | .99 | .99 | .51 |
| DTF (Outflow) | $t_5$ | −4.14 | −2.97 | −6.77 |
| | $p$ | .054 | .188 | .006* |
| | $d$ | 0.7 | 0.5 | 0.3 |
| | $1-\beta$ | .27 | .15 | .09 |
| Alpha Peak | $t_5$ | −0.79 | −0.24 | −0.82 |
| | $p$ | 1 | 1 | 1 |
| | $d$ | 0.3 | 0.0 | 0.3 |
| | $1-\beta$ | .09 | .05 | .09 |
| *Delta Power*$^{+}$ | $t_5$ | −6.94 | −0.35 | −5.44 |
| | $p$ | .006* | 1 | .017* |
| | $d$ | 1.7 | 0.0 | 1.7 |
| | $1-\beta$ | .89 | .05 | .91 |
| *Theta Power*$^{+}$ | $t_5$ | −6.53 | 4.24 | −6.2 |
| | $p$ | .008* | .049* | .01* |
| | $d$ | 1.6 | 0.3 | 2.2 |
| | $1-\beta$ | .87 | .09 | .99 |
| Alpha Power | $t_5$ | −8.61 | −0.93 | −3.33 |
| | $p$ | .002* | 1 | .12 |
| | $d$ | 2.2 | 0.9 | 1.1 |
| | $1-\beta$ | .99 | .4 | .58 |
| *Beta Power*$^{+}$ | $t_5$ | −5.75 | 2.26 | −6.88 |
| | $p$ | .013* | .442 | .006* |
| | $d$ | 1.1 | 0.5 | 1.5 |
| | $1-\beta$ | .57 | .19 | .83 |
| Gamma Power | $t_5$ | 3.09 | 5.86 | −3.75 |
| | $p$ | .163 | .012* | .08 |
| | $d$ | 0.6 | 1.7 | 0.7 |
| | $1-\beta$ | .24 | .89 | .3 |
